# Cumulative Vulnerability and Modern Contraceptive Use in Nigeria: A Latent Class Analysis of the 2018 and 2024 Demographic and Health Surveys

**DOI:** 10.64898/2026.10.05.26364808

**Authors:** Abiodun Y. Omotosho, Adetayo Adetunji

## Abstract

Modern contraceptive use in Nigeria remains low and unequally distributed. Existing research has extensively examined socioeconomic and demographic determinants, but less is known about how multiple dimensions of disadvantage coexist and relate to contraceptive use. This study identified distinct profiles of cumulative vulnerability among Nigerian women, examined inequalities in modern contraceptive use across these profiles, and assessed changes between 2018 and 2024. We analyzed pooled cross-sectional data from the 2018 and 2024 Nigeria Demographic and Health Surveys, comprising 80,871 women aged 15–49 years. Latent class analysis was used to identify vulnerability profiles based on six indicators of socioeconomic, informational and empowerment-related disadvantage. Survey-weighted and multilevel logistic regression models were used to examine associations between vulnerability profiles and modern contraceptive use, including differences by survey year. Average marginal effects and adjusted predicted probabilities were estimated.

Five vulnerability profiles were identified. The extreme vulnerability profile, characterized by predominantly rural residence, limited education, minimal internet and media access, and low decision-making autonomy, was the largest, accounting for 39.2% of women. Its prevalence increased from 38.3% in 2018 to 40.1% in 2024, while the proportion of women in the most advantaged profile increased from 10.4% to 15.7%. Compared with women in the most advantaged profile, those in the extreme vulnerability profile had a 30.9-percentage-point lower adjusted probability of modern contraceptive use (95% confidence interval: −32.8 to −29.0). This difference was 26.5 percentage points (95% confidence interval: −28.4 to −24.7) after accounting for variation between survey clusters. Adjusted contraceptive use increased across all profiles between 2018 and 2024, but substantial inequalities persisted. Evidence of a differential change for the extreme vulnerability profile was present in the survey-weighted model but not in the multilevel model.

Modern contraceptive use in Nigeria is strongly associated with cumulative vulnerability. Despite improvements between 2018 and 2024, substantial inequalities persisted across vulnerability profiles. These findings underscore the importance of addressing overlapping socioeconomic, informational and empowerment-related barriers and monitoring contraceptive-use inequalities across population groups.

## Background

Access to modern contraception is an important public health intervention for improving the health and well-being of women, children, and families [1,2]. Modern contraception contributes to reducing unintended pregnancies, unsafe abortions, and pregnancy-related morbidity and mortality, while improving maternal and child health outcomes [3,1]. Beyond these health benefits, access to contraception can support women’s educational attainment, labour force participation, economic empowerment, and reproductive autonomy, contributing to broader social and economic development [4,2]. Expanding equitable access to modern contraception is therefore a global public health priority, reflected in the Sustainable Development Goals’ commitment to universal access to sexual and reproductive healthcare services by 2030 [5]. Nevertheless, access to and use of modern contraception remain unequally distributed across and within countries [6,7]. Millions of women, particularly in low- and middle-income countries, continue to face barriers to obtaining reproductive health information and services and exercising informed reproductive choice [1, 8]. These global inequalities are particularly relevant to Nigeria, where previous research has documented low modern contraceptive use and persistent unmet need for family planning [9,10]. Recent national estimates indicate that these challenges persist, with substantial geographical and socioeconomic disparities in contraceptive use [11]. Importantly, national contraceptive prevalence estimates conceal considerable heterogeneity across the Nigerian population. According to the 2024 Nigeria Demographic and Health Survey, modern contraceptive prevalence among currently married women ranged from 43% in Ekiti State, 35% in Osun, and 34% in Oyo to just 3% in Kebbi, 4% in Jigawa, and 6% in Sokoto. Substantial geographical disparities were also evident across geopolitical zones, with prevalence highest in the South West (31.4%) and lowest in the North West (9.3%) [11]. Previous studies have documented inequalities associated with educational attainment, household wealth, age, marital status, and place of residence [9, 12,13].

Contraceptive use is also associated with reproductive preferences, women’s decision-making autonomy, partner support, employment status, exposure to mass media, and access to contraceptive services and information [9,14,15]. Community-level conditions, including prevailing social norms, religious and cultural practices, and the availability and accessibility of reproductive healthcare services, may further influence contraceptive decisions [15,16]. Spatial and multilevel research has also documented geographical clustering of modern contraceptive use in Nigeria and associations with community-level literacy and contraceptive knowledge [17].

Women rarely experience socioeconomic, demographic, and contextual disadvantages in isolation. Limited education, poverty, rural residence, and restricted access to health information and services may coexist, creating distinct patterns of vulnerability that are not fully captured when these characteristics are considered separately [18]. These overlapping disadvantages may shape women’s access to contraceptive information and services, their ability to negotiate contraceptive use within relationships, and their capacity to make and act on informed reproductive decisions [19–21]. The Social Determinants of Health framework provides a basis for understanding how unequal access to social, economic, and environmental resources contributes to health inequalities. Intersectionality theory further highlights the importance of considering how social positions and systems of inequality intersect to shape experiences of advantage and disadvantage [22, 18].

Existing research on contraceptive use in Nigeria has established associations with individual and community-level characteristics and documented inequalities across socioeconomic and demographic groups [13,23]. For example, demographic and socioeconomic determinants of modern contraceptive use have been examined [13], alongside individual and community-level factors, including fertility intentions and perceived community support [23]. In this study, vulnerability refers to the co-occurrence of socioeconomic, geographic, informational, and household-level conditions that may constrain women’s access to and use of modern contraception. We conceptualize vulnerability as multidimensional, reflecting how disadvantages across these domains may cluster within women. Although these approaches provide important evidence about the determinants of contraceptive use, they offer more limited insight into how multiple dimensions of vulnerability cluster within women and whether distinct patterns of vulnerability are associated with different levels of contraceptive use. To address this gap, this study uses the 2018 and 2024 Nigeria Demographic and Health Surveys to identify distinct profiles of multidimensional vulnerability among women aged 15–49 years, examine changes in the distribution of these profiles between survey rounds, and assess their associations with modern contraceptive use. It further investigates whether these associations differ by survey year and persist after accounting for variation between survey clusters. The findings aim to inform more equitable and context-responsive family planning programs in Nigeria.

## Methods

### Study Design

This study was a secondary analysis of pooled cross-sectional data from the 2018 and 2024 Nigeria Demographic and Health Surveys (NDHS). Individual recode datasets from both survey rounds were harmonized and pooled to examine patterns of cumulative vulnerability, their associations with modern contraceptive use, and changes between survey years. The study was informed by the Social Determinants of Health framework and Intersectionality Theory, which guided the selection of indicators representing multiple dimensions of social, economic, informational, and empowerment-related vulnerability and the examination of how these dimensions co-occur among women in Nigeria [18, 22].

### Study Population

The study population comprised women aged 15–49 years who completed the women’s questionnaire in the 2018 and 2024 NDHS. The questionnaire was administered to eligible women of reproductive age residing in selected households and collected information on fertility, family planning, reproductive health, maternal and child health, women’s empowerment, and demographic and household characteristics [11]. All interviewed women aged 15–49 years were considered eligible for inclusion, consistent with the study objective of examining patterns of cumulative vulnerability and modern contraceptive use among women of reproductive age in Nigeria.

### Sample size and sampling

The 2018 NDHS included 41,821 women, while the 2024 NDHS included 39,050 women, yielding a pooled sample of 80,871 women. Both surveys employed stratified two-stage cluster sampling to generate nationally representative estimates. In the first stage, enumeration areas were selected within sampling strata. In the second stage, households were systematically selected from the sampled enumeration areas, and eligible women aged 15–49 years in selected households were invited to complete the women’s questionnaire. Sampling weights, cluster identifiers, and stratification variables were retained and harmonized across survey rounds. The latent class analysis included 54,527 women with complete information on the six vulnerability indicators, while the regression analyses included 54,357 women with complete data on all model variables. A total of 170 women in the sterilized respondent or partner category were excluded because this category perfectly predicted modern contraceptive use.

## Study variables

### Outcome Variable

The primary outcome was current modern contraceptive use, derived from the NDHS variable on current contraceptive method. Women reporting use of a modern contraceptive method, including female or male sterilization, oral contraceptive pills, intrauterine devices, injectables, implants, condoms, emergency contraception, and the lactational amenorrhea method, were classified as modern contraceptive users. Women reporting no contraceptive use or use of a traditional method were classified as non-users of modern contraception. The outcome was coded as a binary variable (0 = non-use of modern contraception; 1 = modern contraceptive use).

#### Indicators of cumulative vulnerability

We operationalized cumulative vulnerability using indicators representing socioeconomic, geographic, informational, and household decision-making dimensions. Six indicators of cumulative vulnerability were selected a priori based on the Social Determinants of Health framework, Intersectionality Theory, previous reproductive health research, and comparability across the two NDHS rounds. Educational attainment was categorized as no education, primary, secondary, or higher, while household wealth was categorized into quintiles ranging from poorest to richest. Place of residence was classified as urban or rural. Media exposure was defined as exposure to at least one form of mass media (radio, television, or newspaper), and internet use indicated whether a respondent had used the internet in the preceding 12 months. Women’s decision-making autonomy was categorized as low or moderate/high based on participation in decisions concerning their own healthcare, major household purchases, and visits to family or relatives. All indicators were harmonized across survey rounds before being used to identify latent vulnerability profiles.

#### Covariates

Age, number of children ever born, fertility preference, and geopolitical region were included as covariates in the multivariable regression analyses. Age and number of children ever born were modeled as continuous variables. Fertility preference was categorized as wanting another child, being undecided, wanting no more children, sterilized (respondent or partner), and declared infecund. The sterilized category was excluded from the regression analyses because it perfectly predicted modern contraceptive use. Geopolitical region was categorized as North West, North East, North Central, South East, South South, or South West. These covariates were selected based on their conceptual relevance and established associations with contraceptive use.

## Data analysis

Descriptive statistics were used to summarize respondents’ characteristics overall and by modern contraceptive use status. Continuous variables were compared using survey-adjusted linear regression, while categorical variables were compared using design-adjusted Rao–Scott chi-square tests. Descriptive analyses used the full pooled sample of 80,871 women, with available observations for each variable. Analyses accounted for sampling weights, clustering, and stratification, except where otherwise specified.

Latent class analysis (LCA) was conducted using the *poLCA* package in R to identify patterns of multidimensional vulnerability based on women’s education, household wealth, place of residence, internet use, media exposure, and decision-making autonomy [24]. Assuming conditional independence of the observed indicators within each latent class, the probability of an observed response pattern was expressed as:

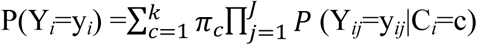

where *i* indexes individual women, *j* indexes the six observed vulnerability indicators, J is the total number of observed indicators (J = 6), K is the number of latent classes, *π_c_* is the probability of membership in class c, and P(Y*_ij_*=y*_ij_*|C*_i_*=c) is the probability of a particular response to indicator *j*, conditional on membership in class c.

Models containing two to six classes were estimated using 20 random starts to reduce the risk of convergence to local maxima, with a maximum of 5,000 iterations per estimation attempt. Model selection considered the Akaike information criterion (AIC), Bayesian information criterion (BIC), entropy, class size, and substantive interpretability. Although the six-class model had lower AIC and BIC, the five-class solution was retained for its higher entropy (0.735 versus 0.698), larger minimum assigned class size (7.0% versus 5.2%), and interpretability. Women were assigned to the class with the highest posterior membership probability. Classification-error-adjusted three-step and six-class sensitivity analyses were conducted to assess the robustness of the findings. The LCA was estimated without survey weights because poLCA does not natively support them. Survey weights were subsequently applied to profile descriptions and regression analyses. Differences in profile distributions between 2018 and 2024 were assessed using a survey-adjusted chi-square test. The potential influence of unweighted LCA on class formation and proportions was acknowledged as a limitation.

Four regression models were estimated to examine associations between vulnerability profiles and modern contraceptive use. Model 1, the primary analysis, used survey-weighted logistic regression accounting for sampling weights, clustering, and stratification. Model 2 extended Model 1 by including an interaction between vulnerability profile and survey year to assess whether associations differed between 2018 and 2024. Models 3 and 4 were weighted multilevel logistic regression models with random intercepts for survey clusters to account for within-cluster correlation. Model 4 additionally included the interaction between vulnerability profile and survey year. The general model specification was expressed as follows:

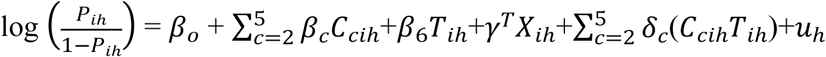

where *P_i_*_ℎ_ is the probability of modern contraceptive use for woman *i* in survey cluster *h*; *C_ci_*_ℎ_ represents vulnerability-profile membership, with the Class 1 (elite advantaged profile) as the reference; *T_i_*_ℎ_ represents survey year; and *X_i_*_ℎ_ includes age, children ever born, fertility preference, and geopolitical region. The coefficients *δ_c_* represent the profile-by-year interactions, and *u*_ℎ_ is the cluster-level random intercept. The interaction terms were omitted from Models 1 and 3, while the random intercept was omitted from Models 1 and 2.

Individual sampling weights were normalized to have a mean of one in the pooled analytical sample and incorporated into the weighted model likelihood. Separate individual- and cluster-level sampling weights were not used; therefore, a multilevel survey-weight scaling procedure using distinct weights at each level was not implemented. Model-based standard errors were used for the multilevel models. The implications of this weighting approach are considered in the study limitations, given that weight scaling can influence estimates from multilevel models fitted to complex survey data [25]. All four models adjusted for survey year, age, children ever born, fertility preference, and geopolitical region. Average marginal effects (AMEs) and their 95% confidence intervals were estimated and expressed as percentage-point differences. For Models 2 and 4, adjusted predicted probabilities of modern contraceptive use were estimated for each vulnerability profile in 2018 and 2024. Profile-by-year interaction coefficients were examined on the log-odds scale and reported in the supplementary material. Joint tests of the profile-by-year interaction were also conducted. Statistical significance was assessed at p < 0.05.

Two sensitivity analyses assessed the robustness of the findings. First, a classification-error-adjusted three-step maximum likelihood approach [26] accounted for uncertainty in class assignment. The estimated classification-error matrix was incorporated into a survey-weighted model with the same covariates as Model 1. Corrected average marginal effects (AMEs) were estimated relative to the elite advantaged profile, with 95% percentile confidence intervals obtained from 500 stratified, rescaled primary sampling unit (PSU) bootstrap replications using the Rao–Wu procedure [27], consistent with applied practice in complex-survey epidemiologic analysis [28]. The procedure treated the LCA solution and classification-error matrix as fixed. Second, the six-class solution was examined using survey-weighted logistic regression with the same covariates. Adjusted predicted probabilities and AMEs were calculated for each of the six profiles, with survey-design-based delta-method 95% confidence intervals. The six-class results were examined to assess whether the associations observed in the primary analysis persisted under an alternative latent class structure.

Data management and descriptive analyses were conducted in Stata/SE 18.0 [29]. Latent class analysis, regression modeling, and sensitivity analyses were conducted in R version 4.6.0 [30] using the *poLCA* (version 1.6.0.2), *survey* (version 4.5), and *lme4* (version 2.0.6) packages, alongside custom routines for classification-error adjustment and bootstrap estimation.

### Ethics statement

The 2018 and 2024 NDHS were conducted with approval from the National Health Research Ethics Committee of Nigeria, with written informed consent obtained from all respondents. This secondary analysis was approved by the Covenant Health Research Ethics Committee (CHREC), Covenant University, Nigeria (Protocol Number CHREC/1457/2026).

## Results

### Respondent characteristics by modern contraceptive use

Overall, 15.8% of women reported current use of a modern contraceptive method, with significant differences across all sociodemographic characteristics (all *p* < 0.001). Compared with non-users, modern contraceptive users were older and had more children ever born. Use increased with educational attainment and household wealth, ranging from 5.7% among women with no formal education to 30.9% among those with higher education, and from 4.7% in the poorest wealth quintile to 27.2% in the richest. Modern contraceptive use was higher among urban (21.6%) than rural (10.6%) women and varied across geopolitical zones, from 9.6% in the North East to 30.4% in the South West. Contraceptive use was also higher among women with internet access (28.6%) compared with those without (12.4%), mass media exposure (20.7% vs. 7.0%), and moderate/high decision-making autonomy (26.0% vs. 10.5%). Among women who wanted no additional children, 27.7% reported modern contraceptive use. Overall, modern contraceptive prevalence increased from 14.3% in 2018 to 17.3% in 2024.

### Characteristics of the latent vulnerability profiles

A five-class solution was selected based on a balance of model fit, classification quality, class sizes and substantive interpretability. Although the six-class model had lower AIC and BIC values, the five-class model had higher entropy (0.735 compared with 0.698) and a larger minimum assigned class proportion (7.0% compared with 5.2%). In the retained five-class solution, the survey-weighted pooled prevalence of the profiles ranged from 7.4% for Class 4 to 39.2% for Class 5 (Table 2). The classes were ordered according to their overall patterns of cumulative disadvantage as shown in Table 2. Class 1 (Elite advantaged) accounted for 12.9% of women and was characterized by high educational attainment, household wealth, internet use, and media exposure. Class 2 (Urban emerging advantage) represented 24.3% and comprised relatively advantaged women, most of whom had secondary education and media exposure but substantially lower internet use than Class 1. Class 3 (Rural transitional) accounted for 16.2% and consisted predominantly of rural women with intermediate educational attainment, moderate media exposure, and limited internet use. Class 4 (Disempowered urban poor) represented 7.4% of women and was predominantly urban, with very low educational attainment and internet use and a high prevalence of low decision-making autonomy. Class 5 (Extreme vulnerability) was the largest profile (39.2%) and was characterized by predominantly rural residence, minimal educational attainment and internet use, limited media exposure, low decision-making autonomy, and concentrated socioeconomic disadvantage.

**Table 1:**
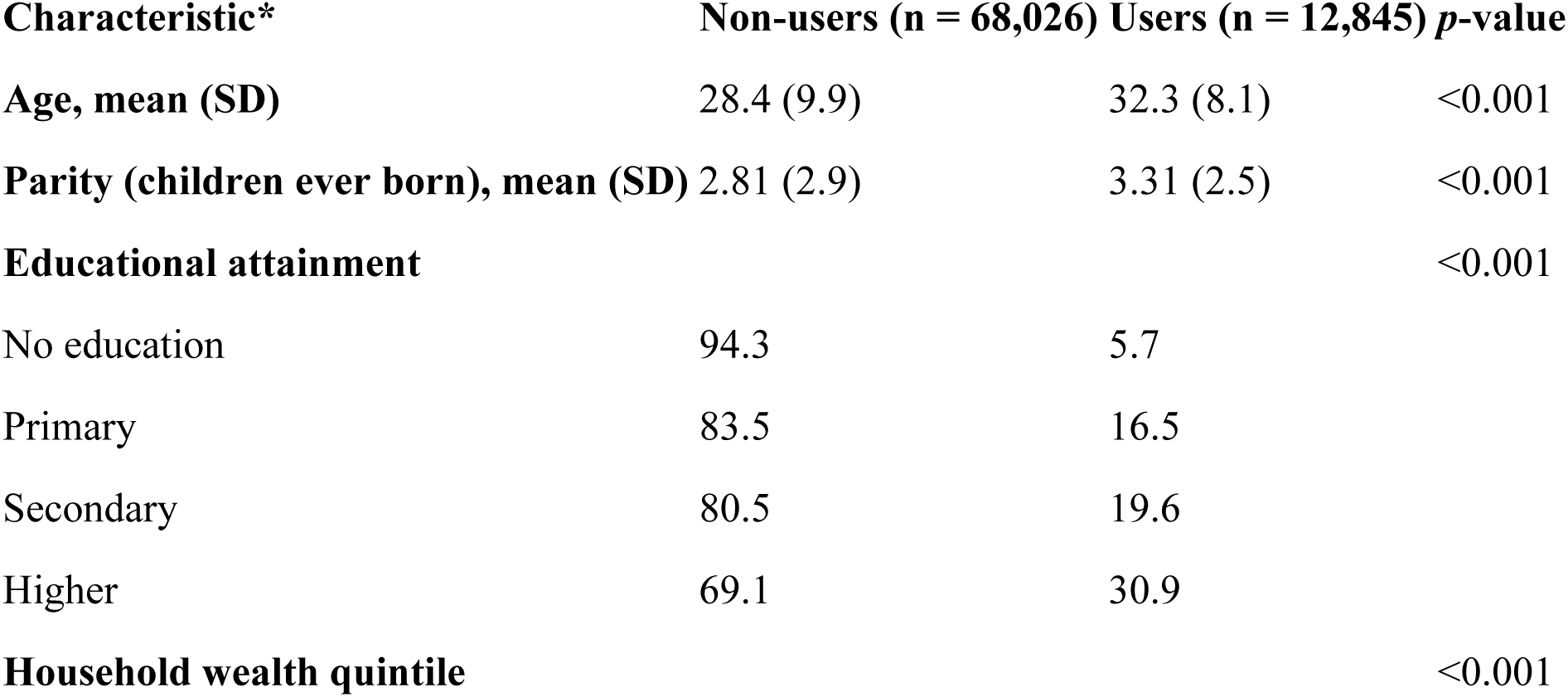
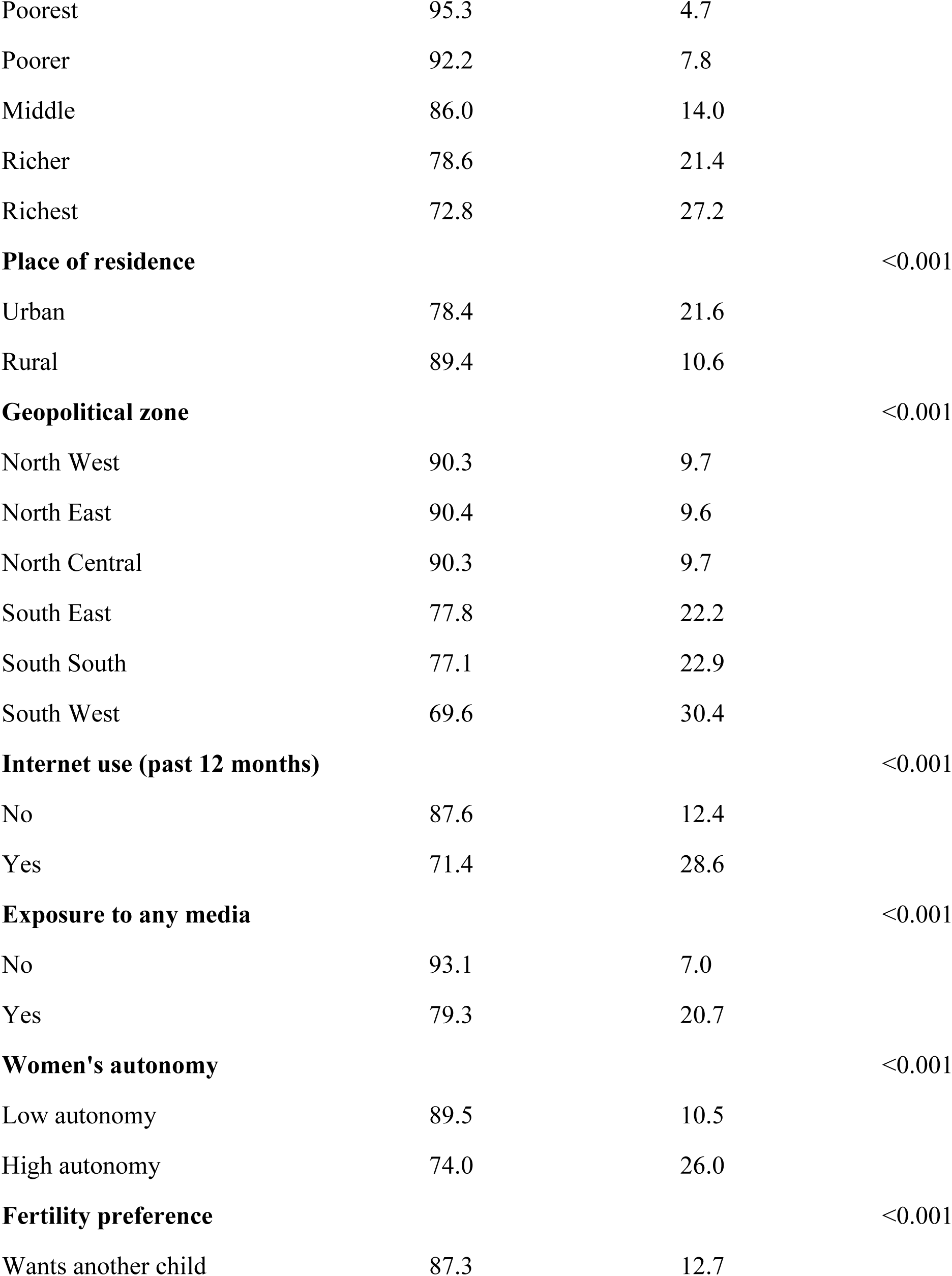
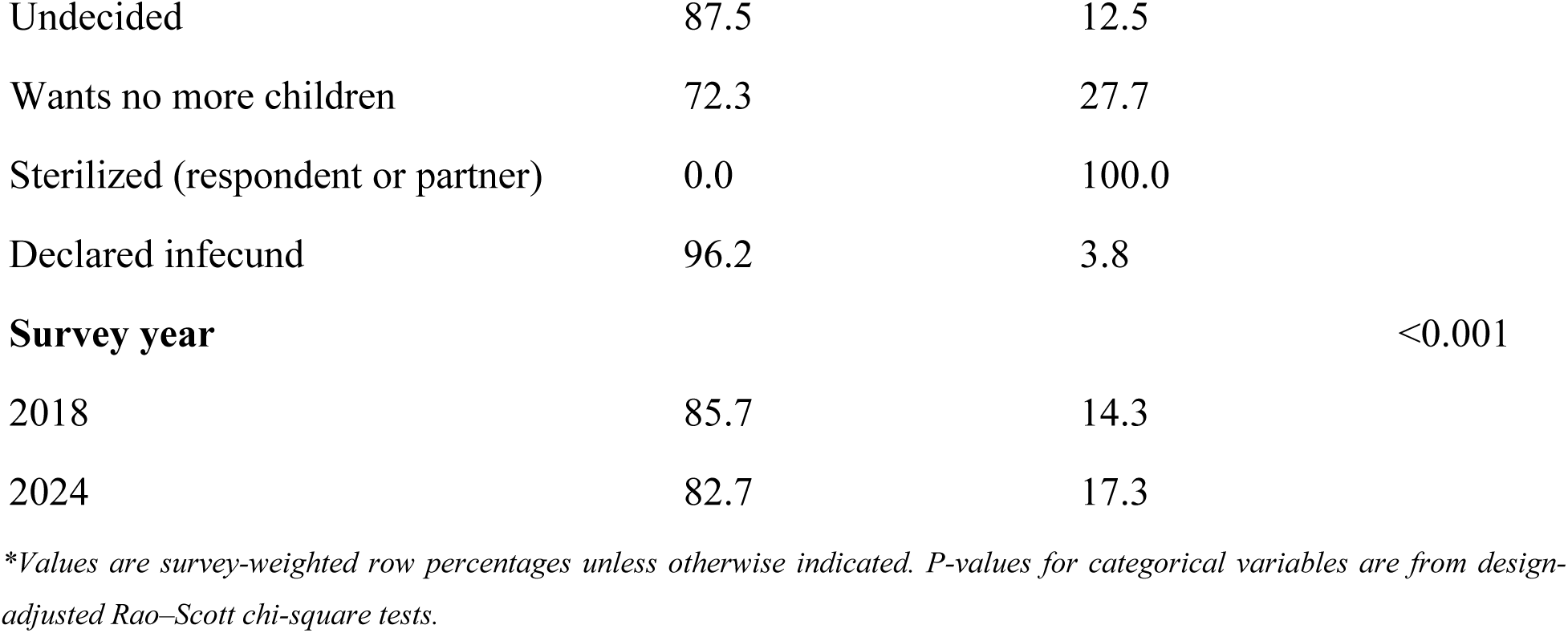
Weighted characteristics of respondents by current modern contraceptive use.

| Characteristic* | Non-users (n = 68,026) | Users (n = 12,845) | p-value |
| --- | --- | --- | --- |
| **Age, mean (SD)** | 28.4 (9.9) | 32.3 (8.1) | <0.001 |
| **Parity (children ever born), mean (SD)** | 2.81 (2.9) | 3.31 (2.5) | <0.001 |
| **Educational attainment** |  |  | <0.001 |
| No education | 94.3 | 5.7 |  |
| Primary | 83.5 | 16.5 |  |
| Secondary | 80.5 | 19.6 |  |
| Higher | 69.1 | 30.9 |  |
| **Household wealth quintile** |  |  | <0.001 |

| **Characteristic\*** | **Non-users (n = 68,026)** | **Users (n = 12,845)** | **p-value** |
| --- | --- | --- | --- |
| Poorest | 95.3 | 4.7 |  |
| Poorer | 92.2 | 7.8 |  |
| Middle | 86.0 | 14.0 |  |
| Richer | 78.6 | 21.4 |  |
| Richest | 72.8 | 27.2 |  |
| **Place of residence** |  |  | <0.001 |
| Urban | 78.4 | 21.6 |  |
| Rural | 89.4 | 10.6 |  |
| **Geopolitical zone** |  |  | <0.001 |
| North West | 90.3 | 9.7 |  |
| North East | 90.4 | 9.6 |  |
| North Central | 90.3 | 9.7 |  |
| South East | 77.8 | 22.2 |  |
| South South | 77.1 | 22.9 |  |
| South West | 69.6 | 30.4 |  |
| **Internet use (past 12 months)** |  |  | <0.001 |
| No | 87.6 | 12.4 |  |
| Yes | 71.4 | 28.6 |  |
| **Exposure to any media** |  |  | <0.001 |
| No | 93.1 | 7.0 |  |
| Yes | 79.3 | 20.7 |  |
| **Women's autonomy** |  |  | <0.001 |
| Low autonomy | 89.5 | 10.5 |  |
| High autonomy | 74.0 | 26.0 |  |
| **Fertility preference** |  |  | <0.001 |
| Wants another child | 87.3 | 12.7 |  |
| Undecided | 87.5 | 12.5 |  |
| Wants no more children | 72.3 | 27.7 |  |
| Sterilized (respondent or partner) | 0.0 | 100.0 |  |
| Declared infecund | 96.2 | 3.8 |  |
| Survey year |  |  | <0.001 |
| 2018 | 85.7 | 14.3 |  |
| 2024 | 82.7 | 17.3 |  |

\*Values are survey-weighted row percentages unless otherwise indicated. P-values for categorical variables are from design-adjusted Rao–Scott chi-square tests.

**Table 2:**
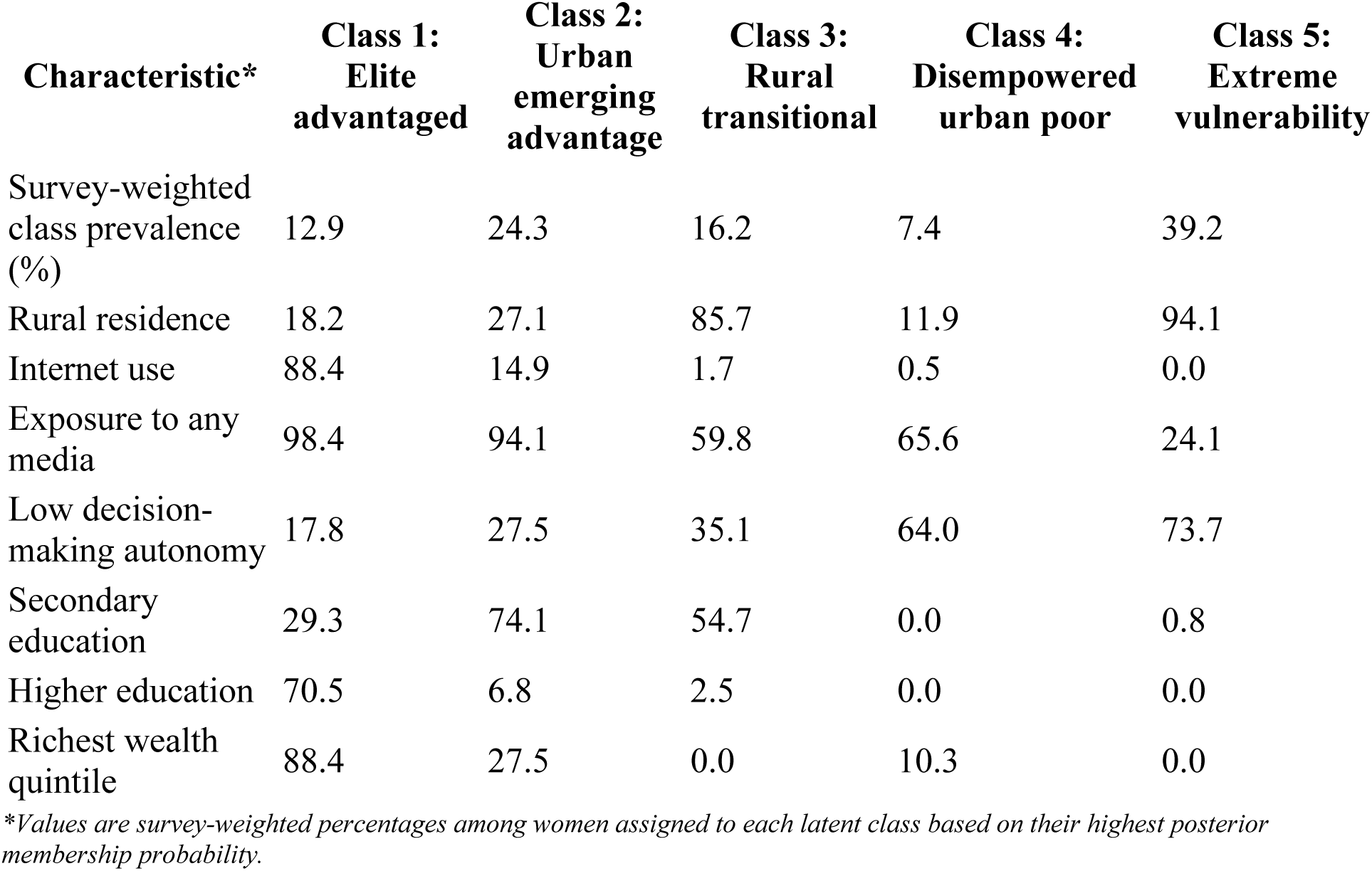
Characteristics of the five latent vulnerability profiles.

### Distribution of latent vulnerability profiles across survey years

The distribution of latent vulnerability profiles differed significantly between 2018 and 2024 (*p* < 0.001). The prevalence of Class 1 increased from 10.4% in 2018 to 15.7% in 2024, representing the largest increase among the five profiles. In contrast, the prevalence of Classes 2, 3, and 4 declined from 25.9% to 22.5%, 17.0% to 15.3%, and 8.4% to 6.4%, respectively. Class 5 remained the largest profile in both survey years, increasing slightly from 38.3% in 2018 to 40.1%.

### Association between vulnerability profiles and modern contraceptive use

Four regression models examined the association between vulnerability profiles and modern contraceptive use (Table 3). In the primary survey-weighted model (Model 1), women in the other four vulnerability profiles had significantly lower adjusted probabilities of modern contraceptive use than women in Class 1, with average marginal effects ranging from −8.6 to −30.9 percentage points. The differences increased across profiles, from −8.6 percentage points (pp; 95% CI: −10.5 to −6.7) for Class 2 to −18.3 pp (95% CI: −20.4 to −16.2) for Class 3, −24.5 pp (95% CI: −27.1 to −21.9) for Class 4, and −30.9 pp (95% CI: −32.8 to −29.0) for Class 5.

**Table 3:** Average marginal effects (percentage-point differences) for the association between latent vulnerability profiles and modern contraceptive use.

| Variable | Model 1<br>AME,<br>pp (95%<br>CI) | p | Model 2<br>AME,<br>pp (95%<br>CI) | p | Model 3<br>AME,<br>pp (95%<br>CI) | p | Model 4<br>AME,<br>pp (95%<br>CI) | p |
| --- | --- | --- | --- | --- | --- | --- | --- | --- |
| **Latent**<br>**vulnerability**<br>**profile (ref:**<br>**Class 1)** |  |  |  |  |  |  |  |  |
| Class 2: Urban<br>emerging<br>advantage | –8.60<br>(–10.52,<br>–6.69) | <0.001 | –8.70<br>(–10.68,<br>–6.71) | <0.001 | –6.85<br>(–8.48,<br>–5.21) | <0.001 | –6.81<br>(–8.49,<br>–5.13) | <0.001 |
| Class 3: Rural transitional | -18.27<br>(-20.39,<br>-16.16) | <0.001 | -18.37<br>(-20.56,<br>-16.19) | <0.001 | -15.76<br>(-17.63,<br>-13.89) | <0.001 | -15.78<br>(-17.69,<br>-13.86) | <0.001 |
| Class 4: Disempowered urban poor | -24.49<br>(-27.10,<br>-21.88) | <0.001 | -24.53<br>(-27.19,<br>-21.87) | <0.001 | -20.28<br>(-22.60,<br>-17.95) | <0.001 | -20.22<br>(-22.58,<br>-17.86) | <0.001 |
| Class 5: Extreme vulnerability | -30.89<br>(-32.82,<br>-28.97) | <0.001 | -31.02<br>(-33.01,<br>-29.02) | <0.001 | -26.54<br>(-28.43,<br>-24.66) | <0.001 | -26.50<br>(-28.44,<br>-24.57) | <0.001 |
| **Survey year (ref: 2018)** |  |  |  |  |  |  |  |  |
| 2024 | 3.44<br>(2.45,<br>4.43) | <0.001 | 3.47<br>(2.48,<br>4.46) | <0.001 | 4.74<br>(3.74,<br>5.73) | <0.001 | 4.77<br>(3.78,<br>5.76) | <0.001 |
| **Continuous covariates** |  |  |  |  |  |  |  |  |
| Age | -0.31<br>(-0.38,<br>-0.24) | <0.001 | -0.31<br>(-0.38,<br>-0.25) | <0.001 | -0.41<br>(-0.47,<br>-0.34) | <0.001 | -0.41<br>(-0.47,<br>-0.34) | <0.001 |
| Children ever born | 1.33<br>(1.12,<br>1.53) | <0.001 | 1.34<br>(1.13,<br>1.54) | <0.001 | 1.76<br>(1.56,<br>1.97) | <0.001 | 1.76<br>(1.56,<br>1.97) | <0.001 |
| **Fertility preference (ref: wants another)** |  |  |  |  |  |  |  |  |
| Undecided | 2.13<br>(0.33,<br>3.94) | 0.021 | 2.04<br>(0.25,<br>3.84) | 0.026 | 0.89<br>(-0.78,<br>2.56) | 0.297 | 0.86<br>(-0.81,<br>2.53) | 0.312 |
| Wants no more | 10.29<br>(9.12,<br>11.46) | <0.001 | 10.31<br>(9.14,<br>11.48) | <0.001 | 9.40<br>(8.24,<br>10.56) | <0.001 | 9.42<br>(8.25,<br>10.58) | <0.001 |
| Declared infecund | -6.63<br>(-8.82,<br>-4.44) | <0.001 | -6.65<br>(-8.84,<br>-4.47) | <0.001 | -6.78<br>(-8.93,<br>-4.62) | <0.001 | -6.78<br>(-8.94,<br>-4.63) | <0.001 |
| **Region (ref: North West)** |  |  |  |  |  |  |  |  |
| North East | 2.63<br>(0.91,<br>4.35) | 0.003 | 2.88<br>(1.16,<br>4.61) | 0.001 | 2.42<br>(0.59,<br>4.26) | 0.010 | 2.61<br>(0.77,<br>4.45) | 0.005 |
| North Central | -0.07<br>(-1.58,<br>1.45) | 0.932 | 0.43<br>(-1.10,<br>1.96) | 0.579 | -0.87<br>(-2.40,<br>0.67) | 0.267 | -0.57<br>(-2.13,<br>1.00) | 0.479 |
| South East | 4.61<br>(3.00,<br>6.22) | <0.001 | 4.71<br>(3.11,<br>6.32) | <0.001 | 5.48<br>(3.66,<br>7.30) | <0.001 | 5.61<br>(3.80,<br>7.42) | <0.001 |
| South South | 2.23<br>(0.74,<br>3.73) | 0.003 | 2.43<br>(0.95,<br>3.92) | 0.001 | 3.82<br>(2.18,<br>5.46) | <0.001 | 3.93<br>(2.29,<br>5.57) | <0.001 |
| South West | 10.83<br>(8.97,<br>12.69) | <0.001 | 10.99<br>(9.15,<br>12.82) | <0.001 | 12.68<br>(10.86,<br>14.50) | <0.001 | 12.79<br>(10.98,<br>14.60) | <0.001 |

Models 2 and 4 included interactions between vulnerability profile and survey year. For these models, profile AMEs were averaged over the observed survey-year distribution, while the survey-year AME was averaged across vulnerability profiles according to their observed distribution. The associations between vulnerability profiles and modern contraceptive use remained statistically significant across all four models, although their magnitudes were attenuated after accounting for survey-cluster random intercepts. In Model 2, profile differences relative to Class 1 ranged from −8.7 pp for Class 2 to −31.0 pp for Class 5. After accounting for survey-cluster random intercepts in Model 4, the corresponding differences were attenuated to −6.8 pp and −26.5 pp, respectively. In Model 3, adjusted differences relative to Class 1 were −6.9 pp (95% CI: −8.5 to −5.2) for Class 2, −15.8 pp (95% CI: −17.6 to −13.9) for Class 3, −20.3 pp (95% CI: −22.6 to −18.0) for Class 4, and −26.5 pp (95% CI: −28.4 to −24.7) for Class 5. The inclusion of profile-by-year interactions in Models 2 and 4 produced similar overall patterns.

After adjustment, modern contraceptive use was higher in 2024 than in 2018, with average marginal effects of 3.4 pp (95% CI: 2.5 to 4.4) in Model 1 and 4.7 pp (95% CI: 3.7 to 5.7) in Model 3. Across the models, older age was associated with lower modern contraceptive use, whereas a greater number of children ever born and wanting no additional children were associated with higher use. Geographic differences in modern contraceptive use persisted after adjustment for other factors. Women in the South West had the largest positive difference compared with those in the North West, with an adjusted probability of modern contraceptive use that was 10.8 percentage points higher in Model 1 and 12.7 percentage points higher in Model 3.

### Variation in associations across survey years

The joint test of the profile-by-year interaction was statistically significant in the survey-weighted model (Model 2; design-adjusted F(4, 2600) = 2.93, p = 0.020), providing evidence of variation in the associations between vulnerability profiles and modern contraceptive use across survey years. The corresponding joint test in the multilevel model (Model 4) did not reach conventional statistical significance (Wald χ²(4) = 8.94, p = 0.063). Adjusted predicted probabilities increased between 2018 and 2024 across all five profiles in both interaction models (Table 4). In Model 2, predicted modern contraceptive use increased from 33.9% to 39.7% for Class 1 and from 4.3% to 7.1% for Class 5. In Model 4, the corresponding probabilities increased from 29.6% to 37.5% for Class 1 and from 5.1% to 8.2% for Class 5. Although contraceptive use increased across all profiles, the gap between Class 1 and Class 5 remained substantial in both survey years.

**Table 4:**
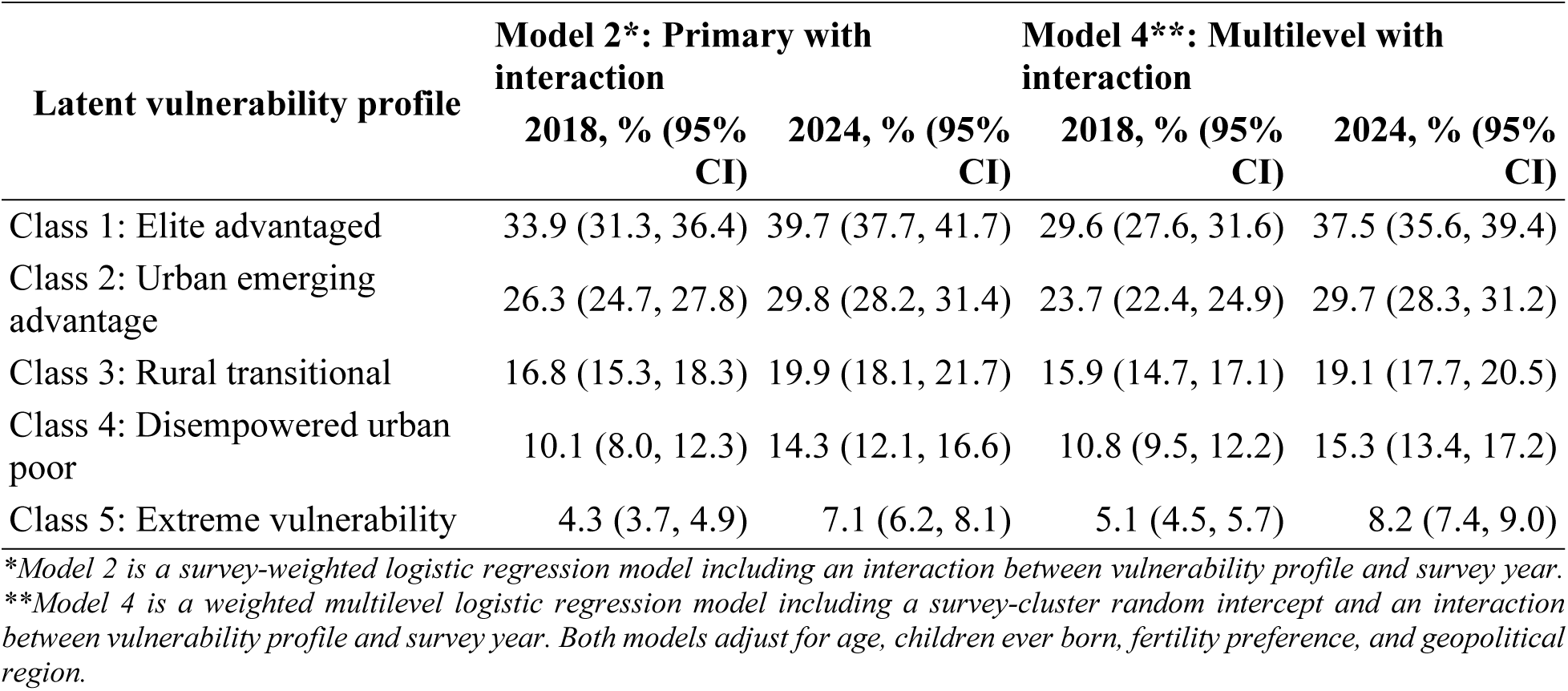
Adjusted predicted probabilities (%) of modern contraceptive use by latent vulnerability profile and survey year.

### Sensitivity analyses

Two sensitivity analyses were conducted to assess the robustness of the associations between vulnerability profiles and modern contraceptive use (Supplementary Table 8). The classification-error-adjusted three-step analysis produced associations consistent in direction with the primary survey-weighted regression model (Model 1), although the estimated differences were generally larger. Compared with Class 1, the adjusted differences in modern contraceptive use were −8.4 percentage points (pp; 95% CI: −11.0 to −6.0) for Class 2, −21.7 pp (95% CI: −24.1 to −18.6) for Class 3, −30.9 pp (95% CI: −34.2 to −27.2) for Class 4, and −36.1 pp (95% CI: −38.6 to −33.8) for Class 5. These findings indicate that accounting for classification error did not alter the direction or overall pattern of the observed associations. The alternative six-class solution similarly showed substantial differences in modern contraceptive use across vulnerability profiles. Using Class 1 (Elite urban advantage) of the six-class solution as the reference, the adjusted differences were −19.3 pp (95% CI: −21.4 to −17.2) for Class 2, −25.5 pp (95% CI: −27.8 to −23.1) for Class 3, −8.7 pp (95% CI: −10.7 to −6.7) for Class 4 (Established urban advantage), −9.5 pp (95% CI: −12.2 to −6.9) for Class 5, and −31.3 pp (95% CI: −33.2 to −29.4) for Class 6 (Extreme rural vulnerability). Although the six-class solution distinguished additional socioeconomic profiles, substantial differences in modern contraceptive use across vulnerability profiles remained evident.

## Discussion

This study identified five distinct vulnerability profiles among Nigerian women, revealing substantial inequalities in modern contraceptive use associated with different combinations of socioeconomic, informational, geographic, and empowerment-related characteristics. Women in the extreme vulnerability profile had substantially lower adjusted probabilities of modern contraceptive use than those in the elite advantaged profile. These associations persisted across survey-weighted and multilevel regression models and were also evident in analyses accounting for uncertainty in latent class assignment and using an alternative six-class solution. Although predicted modern contraceptive use increased across all profiles between 2018 and 2024, substantial inequalities remained.

The vulnerability profiles identified in this study demonstrate how social and economic disadvantages cluster among Nigerian women. Previous studies in Nigeria and sub-Saharan Africa have established associations between modern contraceptive use and individual factors such as education, household wealth, residence, access to information, and women’s empowerment [12, 17, 31]. Our findings extend this evidence by showing how these disadvantages coexist within distinct vulnerability profiles rather than occurring in isolation. From a Social Determinants of Health perspective, such overlapping disadvantages are important because barriers across multiple domains may reinforce one another [32,18]. For example, limited education, combined with economic deprivation, restricted access to information, and low decision-making autonomy, may constrain women’s opportunities to obtain reproductive health information, access services, and make informed contraceptive decisions.

We found that the distribution of the identified profiles highlights the persistence of cumulative vulnerabilities. While the proportion of women in the elite advantaged profile increased between 2018 and 2024, the extreme vulnerability remained the largest profile and grew slightly. This persistence may reflect structural barriers that limit opportunities for women experiencing multiple disadvantages. Research in Nigeria has shown that inadequate rural infrastructure and gendered unpaid care responsibilities constrain women’s opportunities for education and income-generating activities [33], while social empowerment is associated with poverty reduction among women in agriculture [34]. These longstanding constraints may have been compounded by economic shocks during the study period. The COVID-19 pandemic disrupted women’s informal livelihoods and increased their unpaid care responsibilities, with experiences shaped by existing socioeconomic inequalities [35]. This evidence suggests that overlapping disadvantages may limit women’s capacity to withstand economic shocks and improve their socioeconomic circumstances.

Modern contraceptive use was substantially lower among women experiencing cumulative vulnerability, with the largest adjusted difference between the extreme vulnerability and elite advantaged profiles. This is consistent with Nigerian studies showing higher contraceptive use among women with greater educational and economic resources, more participation in healthcare decisions, and greater exposure to mass media [36, 17]. Use was also considerably lower in the disempowered urban poor and rural transitional profiles, indicating that disadvantage does not have to be extreme to suppress contraceptive use. The findings among the urban poor are consistent with evidence from six Nigerian cities showing that the apparent urban advantage can conceal low contraceptive use among poorer women [37]. Limited autonomy may represent another constraint, as sexual autonomy remained associated with modern contraceptive use after adjustment for education, wealth, and other socioeconomic characteristics [38].

The associations between vulnerability profiles and modern contraceptive use remained substantial in the multilevel models, although the estimated differences were somewhat smaller than in the primary analysis. One possible explanation is that the multilevel models account for unobserved conditions shared by women within the same survey clusters. Such conditions could include differences in local service environments or community norms, although these factors were not directly measured at the cluster level in this analysis. Evidence from Nigeria demonstrates that perceived community support for family planning and awareness of contraceptive use among friends and relatives are associated with modern contraceptive use [23]. Similarly, changes in gender-equitable attitudes toward household decision-making and couples’ family planning decisions at both individual and neighborhood levels were associated with modern contraceptive adoption and continuation among women in urban Nigeria [39].

The sensitivity analyses reinforced the main findings, with substantial inequalities in modern contraceptive use persisting after accounting for classification error and under an alternative six-class solution. The larger estimated differences following classification-error correction suggest that modal class assignment may have attenuated the associations observed in the primary analysis. Although the magnitude of these associations varied across analytical approaches, the overall pattern of lower contraceptive use among women experiencing cumulative disadvantage remained consistent.

Modern contraceptive use increased across all vulnerability profiles between 2018 and 2024, but substantial disparities persisted between the most and least advantaged women. This finding is consistent with [12], who documented increasing contraceptive use in Nigeria alongside persistent socioeconomic inequalities. Women with greater financial and informational resources may be better positioned to benefit from improvements in family planning services, whereas those experiencing cumulative disadvantage may face multiple barriers that limit such benefits. Nigerian studies have documented inequalities associated with household wealth, education, residence, and decision-making autonomy [40, 41]. Qualitative research has also identified overlapping barriers, including spousal opposition, concerns about side effects, difficulties reaching facilities, and inadequate contraceptive supplies [42,43]. Although the first interaction model (Model 2) indicated significant differences in changes in contraceptive use across vulnerability profiles, the interaction was not statistically significant in the multilevel model (Model 4). The pattern of predicted probabilities nevertheless showed uneven gains across profiles, with the largest increase observed among women in the most advantaged profile and generally smaller gains among more vulnerable profiles. However, the evidence that these changes differed systematically across profiles was weaker after accounting for differences between communities. What remained consistent was the substantial gap in contraceptive use between the most advantaged and more disadvantaged profiles.

### Strengths and Limitations

Several limitations should be considered when interpreting these findings. First, the repeated cross-sectional DHS data preclude causal inference, prevent tracking individual changes in vulnerability, and leave open the possibility of reverse causality. Second, self-reported contraceptive use may be subject to reporting bias. Third, the latent profiles were based on available DHS indicators and may not capture other dimensions of vulnerability, including partner dynamics, social norms, and health system barriers. The unweighted latent class analysis may have influenced the estimated class structure. Although subsequent analyses incorporated survey weights and corrected for classification error, the bootstrap procedure treated the LCA solution and classification-error matrix as fixed, leaving some uncertainty unaccounted for. Complete-case analysis may have introduced selection bias, and residual confounding remains possible. Finally, evidence of differential changes in contraceptive use across profiles varied between the primary and multilevel models.

The strengths of this study include its identification of distinct patterns of co-occurring vulnerabilities among Nigerian women, providing insight beyond approaches that treat these factors as separate variables and offer limited information on how they cluster within women. Furthermore, the use of nationally representative data from two DHS rounds enabled comparisons of vulnerability patterns and contraceptive inequalities between 2018 and 2024. Also, the three-step classification-error correction and alternative six-class solution provided important sensitivity checks. The persistence of substantial inequalities across these analyses, as well as the primary and multilevel models, strengthens confidence in the association between cumulative vulnerability and lower modern contraceptive use.

## Conclusion

This study identified five distinct vulnerability profiles among Nigerian women, demonstrating substantial inequalities in modern contraceptive use associated with overlapping socioeconomic, geographic, informational, and empowerment-related disadvantages. Although contraceptive use increased across all profiles between 2018 and 2024, women experiencing the greatest cumulative disadvantage continued to have the lowest probability of use. These findings underscore the persistence of contraceptive inequalities despite improvements in uptake. These findings highlight the importance of considering multiple, co-occurring disadvantages when designing and targeting family planning programs. Efforts to improve equitable contraceptive access may benefit from addressing barriers related to reproductive health information, affordability and availability of quality family planning services, economic disadvantage, and women’s reproductive autonomy.

## Data Availability

No data were generated by this study. The study used publicly available data from the 2018 and 2024 Nigeria Demographic and Health Surveys (NDHS). The datasets are available from the DHS Program upon registration and approval of a data access request at the DHS Program website.

https://dhsprogram.com/data

